# A simple risk score for chronic kidney disease using administrative data: A population-based cohort study

**DOI:** 10.1101/2024.04.09.24305566

**Authors:** Marcello Tonelli, Natasha Wiebe

**Author notes:** **Correspondence to:** Marcello Tonelli, University of Calgary, 7th Floor, CWPH Building, 3280 Hospital Drive NW, Calgary, Alberta, Canada T2N 4Z6.

## Abstract

**Background:** We did this study to develop and validate a risk score for new chronic kidney disease (CKD), focusing on predictors that are typically available in Canadian administrative health datasets.

**Methods:** This was a retrospective population-based cohort study using data from the Alberta Kidney Disease Network database: 3,558,192 adult participants were followed from April 1, 2007 to March 31, 2019. We developed a simple score to predict reduced glomerular filtration rate using bootstrapping (100 iterations with replacement) and internally validated the score using the original dataset.

**Findings:** The final score had a maximum total of 9 points: age 50-70 years, moderate albuminuria, hypertension, diabetes and heart failure all received a single point, and age >70 years and severe albuminuria received three points. The C-statistic of the score for incident CKD was 0.9272 and the Brier score was 0.0053, indicating excellent discrimination. Graphical analysis demonstrated that predicted risk closely aligned with the observed risk of developing CKD, indicating a well-calibrated model.

**Interpretation:** We have derived and internally validated a risk score for new CKD which is suitable for application to routinely collected Canadian administrative health data.

**Funding:** David Freeze chair in health services research

## Background

Chronic kidney disease (CKD) is a common condition that is associated with adverse outcomes and high health costs. CKD can be defined by reduced glomerular filtration rate (eGFR; typically <60 mL/min/1.73m^2^), by increased albuminuria, or by structural and functional abnormalities of the kidneys, although the latter is not commonly used to define CKD in epidemiological studies. Multiple characteristics are associated with excess risk of reduced eGFR, including age, diabetes, hypertension, heart failure and vascular disease, as well as increased albuminuria.

Risk scores have been previously devised for predicting the likelihood of incident CKD, but the best validated of these scores includes multiple characteristics that are not typically available in administrative health datasets.^1^ Other such scores have been validated in relatively small populations, do not include a full range of clinical characteristics that may confer risk, or may not be generalizable to the Canadian population.^2-4^

We did this study to develop and validate a risk score for reduced eGFR (<60 mL/min/1.73m^2^), focusing on predictors that are typically available in Canadian administrative health datasets.

## Methods

We reported this retrospective population-based cohort study according to the STROBE guidelines.^5^ The institutional review boards at the Universities of Calgary and Alberta (REB16-1575/psite00000147) approved the study and waived the requirement for informed consent.

### Data sources and cohort

We used the Alberta Kidney Disease Network (AKDN) database, which incorporates patient registry, provider claims, hospitalizations, ambulatory care utilization, and pharmaceutical information network data from all adults registered with the provincial health ministry in Alberta, Canada; and links them with data from provincial clinical laboratories. This database has been widely used^6-8^ because of its population-based coverage of a geographically defined area, including demographic characteristics, health services utilization, and clinical outcomes. Additional information on the database is available elsewhere, including the validation of selected data elements.^9^ All Alberta residents are eligible for insurance coverage by Alberta Health with >99% participation. We used the database to assemble an open cohort of adults, without CKD, who resided in Alberta, Canada between April 1, 2007 and March 31, 2018. The index date was April 1, 2007, the day of first contact with Alberta Health, or the participant’s 18^th^ birthday, whichever was latest. We followed participants until March 31, 2019, development of CKD, death, or migration from Alberta.

### Chronic kidney disease

We based the definition of CKD on the median outpatient estimated GFR during each fiscal year. We defined CKD as eGFR <60 mL/min/1.73m^2^ (KDIGO stage G3a or higher) and/or registration with a provincial kidney replacement program. We calculated eGFR using the CKD-Epidemiology Collaboration (EPI) equation without race adjustment.^10^

### Risk factors

We selected risk factors for CKD from a list of characteristics that were included in a prediction equation for risk of incident CKD by Nelson *et al*.^*1*^ Of the 8 characteristics in the model (ignoring 2 for eGFR) for people without diabetes, 5 were available in our dataset and were included as potential risk factors in the current manuscript: age, sex, cardiovascular disease, hypertension, and albuminuria. We also considered diabetes as a risk factor for CKD. Ethnicity, smoking status and body mass index were not available in this dataset.

We defined albuminuria using the median outpatient measurement during each fiscal year of any one of the following: albumin: creatinine ratio (ACR), protein:creatinine ratio (PCR), or dipstick urinalysis. We used the PCR assessment when ACR was not available, and used dipstick results when PCR was not available. We categorized measurements as follows: missing, none/mild (ACR <3 mg/mmol, PCR <15 mg/mmol, dipstick negative/trace), moderate (ACR 3-30 mg/mmol, PCR 15-50 mg/mmol, dipstick 1+), and severe (ACR >30 mg/mmol, PCR >50 mg/mmol, dipstick ≥2+).

We defined diabetes, hypertension, heart failure, myocardial infarction, peripheral arterial disease (PAD), and stroke or transient ischemic attack (TIA) using validated algorithms as applied to Canadian provider claims, hospitalizations, and ambulatory care data, each of which had positive predictive values ≥70% as compared to a gold standard measure such as chart review.^11^ We defined coronary artery disease (CAD) by the occurrence of myocardial infarction and/or receipt of a percutaneous coronary intervention or a coronary artery bypass graft. We defined cardiovascular disease as CAD, PAD, and/or stroke/TIA. We included claims data from as far back as April 1994 where records were available.^10^ We considered each participant to have each of these conditions at the beginning of the fiscal year in which it was diagnosed. Detailed methods and the specific algorithms used for comorbidity assessment are available elsewhere.^11^ As in our prior work, we used administrative data to identify age and sex.

### Statistical analyses

We did all analyses using Stata MP 18·0 (www.stata.com). We reported baseline descriptive statistics as counts and percentages, or medians with interquartile ranges.

We followed Sullivan *et al*’s^12^ approach to develop a simple score to predict new CKD. We considered several parametric survival models: log-normal, log-logistic, and Gompertz. Because the model using the log normal distribution had the smallest log-likelihood, we chose that model to regress time to new CKD on the previously selected 7 risk factors: age (<50, 50-70, >70y), sex (male, female), albuminuria (mild/none/missing, moderate, severe), hypertension, diabetes, cardiovascular disease, and heart failure. We developed the score using bootstrapping (100 iterations with replacement of the dataset).^13,14^ We internally validated the score using the original dataset, and plotted the observed risk of incident CKD associated with scores from lowest (0) to highest (9) against the predicted risk of CKD. We reported the Brier score and the C-statistic (the area under the receiver operator curve). The Brier score measures the differences between predicted and observed risk; smaller values indicate less deviance, thus, better calibration. Larger C-statistics indicate better discrimination.

## Results

### Participants

There were 3,558,192 participants in this study with a median of 10 fiscal years of follow-up (interquartile range 8,10; range 1,10). In the last year of follow-up, the median age was 48 years; 51% were male. The population had hypertension (26%), diabetes (10%), cardiovascular disease (8%) and heart failure (3%) (Table 1). Approximately 6% of participants had moderate or severe albuminuria.

**Table 1.** Demographic and clinical characteristics at last follow-up.

|  | All |
| --- | --- |
| N | 3,558,192 (100.0) |
| Age, y | 48 [37,61] |
| Male | 1,818,575 (51.1) |
| Rural residence | 292,792 (9.0) |
| Material deprivation |  |
| 1 (Least deprived) | 661,808 (20.8) |
| 2 | 594,302 (18.6) |
| 3 | 628,606 (19.7) |
| 4 | 640,166 (20.1) |
| 5 (Most deprived) | 663,408 (20.8) |
| Albuminuria |  |
| Mild/none/unmeasured | 3,342,988 (94.0) |
| Moderate | 170,257 (4.8) |
| Severe | 44,947 (1.3) |
| Hypertension | 914,575 (25.7) |
| Diabetes mellitus | 361,618 (10.2) |
| Cardiovascular disease | 268,301 (7.5) |
| Stroke and/or TIA | 175,090 (4.9) |
| Coronary artery disease | 94,141 (2.6) |
| Peripheral artery disease | 33,907 (1.0) |
| Heart failure | 114,676 (3.2) |

IQR interquartile range, TIA transient ischemic attack, y years N (%) or median [IQR]

### Training

Table 2 gives the estimated coefficients and the points assigned using the bootstrapped training approach. Moderate albuminuria was assigned a single point. As a consequence, sex and cardiovascular disease were dropped from the score. Age 50-70 years, hypertension, diabetes and heart failure also received a single point, and age >70 years and severe albuminuria received three points for a maximum total of 9 points.

**Table 2.** Derivation of CKD score.

|  | Value | Coefficient | Points |
| --- | --- | --- | --- |
| Age, y |  |  |  |
| 18-49 | 0 |  | 0 |
| 50-70 | 1 | -0.485024 | 1 |
| >70 | 2 | -1.140609 | 3 |
| Sex |  |  |  |
| Female | 0 |  | 0 |
| Male | 1 | 0.1170866 | 0 |
| Albuminuria |  |  |  |
| Mild/None/Missing | 0 |  | 0 |
| Moderate | 1 | -0.346868 | 1 |
| Severe | 2 | -1.078683 | 3 |
| Hypertension |  |  |  |
| No | 0 |  | 0 |
| Yes | 1 | -0.400835 | 1 |
| Diabetes mellitus |  |  |  |
| No | 0 |  | 0 |
| Yes | 1 | -0.175541 | 1 |
| Cardiovascular disease |  |  |  |
| No | 0 |  | 0 |
| Yes | 1 | -0.157393 | 0 |
| Heart failure |  |  |  |
| No | 0 |  | 0 |
| Yes | 1 | -0.516182 | 1 |
| Constant |  | 11.42099 |  |
| /Sigma |  | 0.4961532 |  |

CKD chronic kidney disease, y years
We used a log-normal survival model to regress CKD onto the preselected risk factors. Sigma is the ancillary parameter of the log-normal model; it represents the standard deviation of the natural logarithm for the CKD variable. The model was trained using the bootstrap (100 iterations with replacement). Moderate albuminuria was selected to have a point of 1. The maximum score is 9. Sex and cardiovascular disease were dropped from the score.

### Validation

The C-statistic of the score for incident CKD was 0.9272 and the Brier score was 0.0053, indicating excellent discrimination. Figure 1 shows that the predicted risk closely aligns with the observed risk of developing CKD, indicating a well-calibrated model. The predicted risk tended to be slightly lower than the observed risk in participants at greater risk for CKD, and slightly higher in those at lower risk for CKD. For example, scores of 0, 3, 5, and 9 were associated with predicted risks of 0.3%, 1.3%, 3.0% and 11.7% respectively, as compared to observed risks of 0.0%, 1.4%, 5.1% and 14.6%. The model remained reasonably well-calibrated in groups defined by age (Supplement Figure S1), sex (Supplement Figure S2), and diabetes status (Supplement Figure S3), although it tended to underestimate risk among younger individuals.

**Figure 1.**
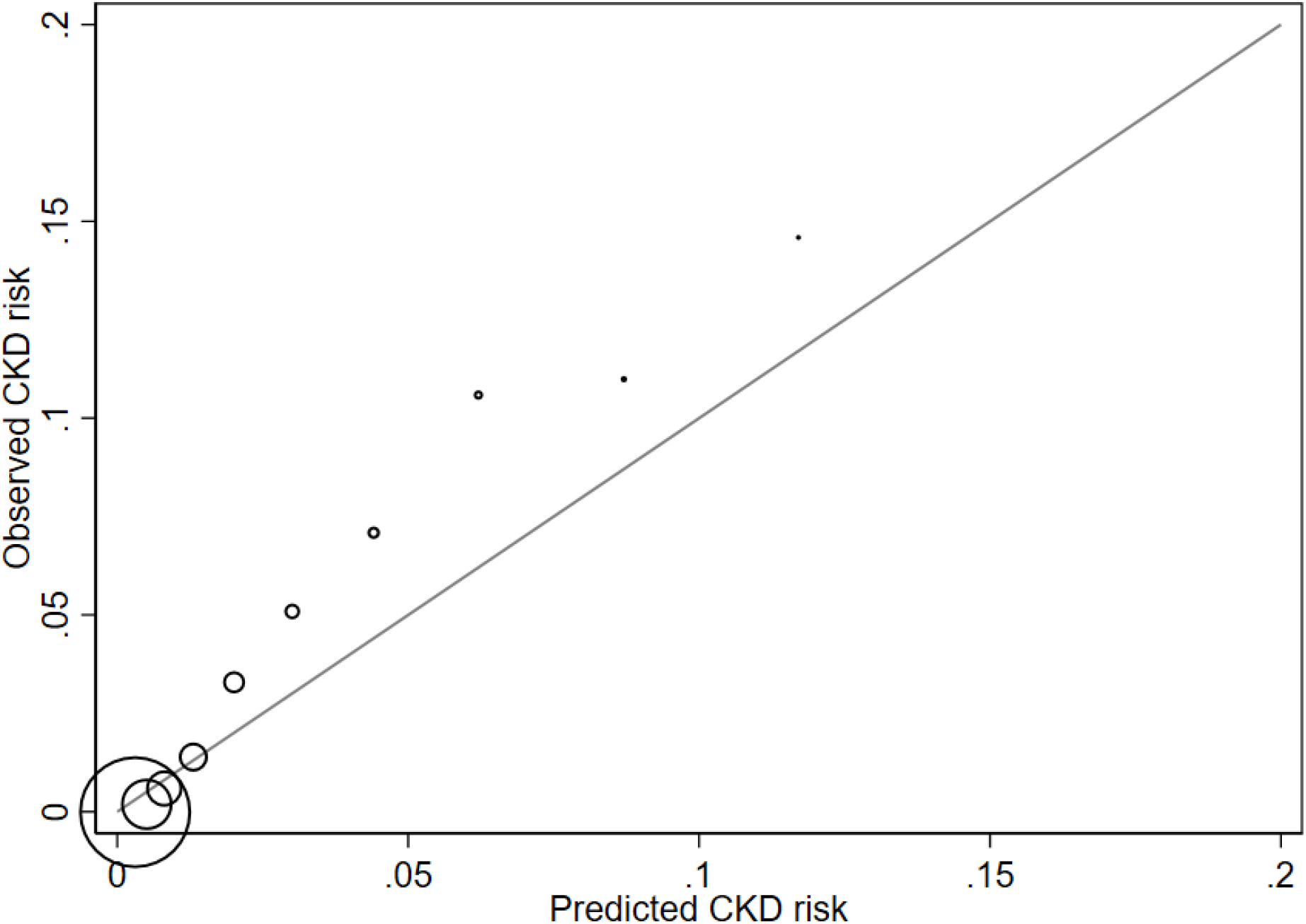
Observed versus predicted risk of CKD. CKD chronic kidney disease The size of the marker reflects the relative number of participants that fall into that risk category.

Figure 2 shows the predicted risk of incident CKD that was associated with combinations of age (<50, 50-70, >70y), albuminuria (none/mild/missing; moderate; severe) and the number of comorbidities (hypertension, diabetes, and/or heart failure).

**Figure 2.**
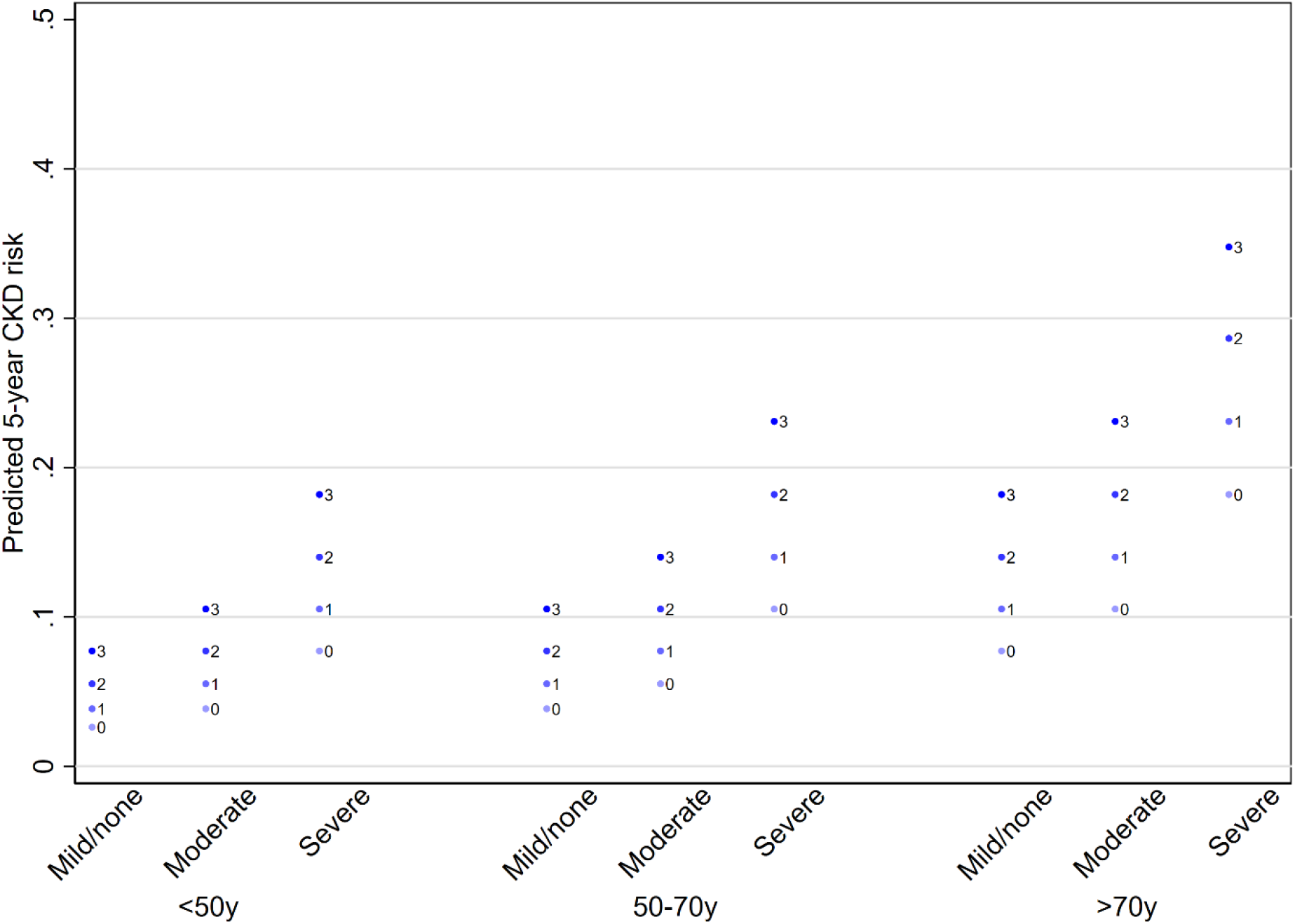
Predicted 5-year risk of CKD by score components. CKD chronic kidney disease Predicted risk is based on 3 categories of age, 3 categories of albuminuria and the number of morbidities (hypertension, diabetes, and/or heart failure; the number of morbidities from 0 to 3 is represented as numerals within each column).

## Discussion

We used data from more than 3.5 million adults treated in a universal health system to derive and internally validate a risk score to predict incident CKD as defined by eGFR <60 mL/min/1.73m^2^, focusing on characteristics that are typically available in Canadian administrative health data while including follow-up of up to 10 years. Our final risk score includes 5 characteristics that can be objectively assessed using routinely available data, and demonstrated excellent calibration with good discrimination overall as well as in key subgroups defined by age, sex and diabetes status. These findings suggest that our score can be appropriately used in combination with Canadian administrative data to predict the risk of incident CKD.

Previous studies have derived and validated risk scores for incident CKD in a variety of populations. The most extensively validated score was generated by the Chronic Kidney Disease Prognosis Consortium using data from more than 5.2 million individuals in 34 cohorts. This score demonstrated excellent discrimination across multiple populations and was derived and validated using rigorous methods. However, it includes several characteristics including smoking status, race/ethnicity and body mass index that are not typically available in Canadian administrative data. Similarly, Lee *et al* derived and validated a risk score for new CKD using data from more than 11 million Korean adults but included multiple parameters that are not available in Canadian administrative data.^15^ O’Seaghda *et al* developed a risk score for incident CKD and validated it in 1777 people aged >45 years from the US population.^3^ Chien *et al* developed an alternative score for incident CKD and validated it in 3205 people aged >35 years from the Chinese population; the score included parameters such as serum urate, body mass index and diastolic blood pressure.^4^ All of these scores are potentially useful for predicting the risk of incident CKD, but none are suitable for use with Canadian administrative health data or were extensively validated in a Canadian population.

Our study has important strengths including its rigorous statistical methods and its relatively large sample size drawn from a population-based sample. However, our study also has limitations that should be considered when interpreting results. First, we considered only CKD as defined by median outpatient estimated GFR<60 mL/min/1.73m^2^, meaning that some participants will have had only a single value of eGFR. Since current definitions of CKD require two values of GFR obtained at least 3 months apart, some participants may have been misclassified with respect to CKD status. In addition, we did not consider CKD as defined by albuminuria or structural kidney abnormalities rather than reduced eGFR. However, our definition is similar to that used in most published studies. Second, despite its excellent discrimination, our score tended to underestimate the true risk of incident CKD at higher values of risk. Similarly, although the score performed well overall, it tended to underestimate risk among younger individuals. However, from a clinical perspective the absolute magnitude of these errors was relatively modest. Third, we did not have access to a suitable dataset for external validation and thus our dataset was only internally validated using bootstrap methodology as well as being derived in a single Canadian province. Future studies could consider validating our score in a sample drawn from a different Canadian jurisdiction, which would help to confirm external validity as well as internal validation.

In conclusion, we have derived and internally validated a risk score for incident CKD as defined by eGFR <60 mL/min/1.73m^2^, and which is suitable for application to routinely collected Canadian administrative health data. This score may be useful to epidemiological and health services researchers.

## Data Availability

We cannot make our dataset available to other researchers due to our contractual arrangements with the provincial health ministry (Alberta Health), who is the data custodian. Researchers may make requests to obtain a similar dataset at https://absporu.ca/research-services/service-application/.

## Authors’ Contributions

MT conceived the study. MT and NW designed the study. NW did the statistical analyses. MT and NW wrote the first draft of the manuscript. All authors contributed to the design, interpretation of results and critical revision of the article for intellectually important content. MT had full access to all the data in the study and takes responsibility for the integrity of the data and the accuracy of the data analysis.

## Acknowledgements

The authors of this report are grateful to Ghenette Houston for administrative support and Ben Vandermeer for his technical review.

## Conflicts of Interest Statement

No conflict of interest to declare.

## Funding

The study was supported by MT’s David Freeze Chair in Health Services Research at the University of Calgary. The sponsors had no role in the design and conduct of the study; collection, management, analysis, and interpretation of the data; preparation, review, or approval of the manuscript; nor in the decision to submit the manuscript for publication.

## Disclaimer

This study is based in part by data provided by Alberta Health and Alberta Health Services. The interpretation and conclusions contained herein are those of the researchers and do not represent the views of the Government of Alberta or Alberta Health Services. Neither the Government of Alberta nor Alberta Health or Alberta Health Services express any opinion in relation to this study.

---

**Figure S1.**
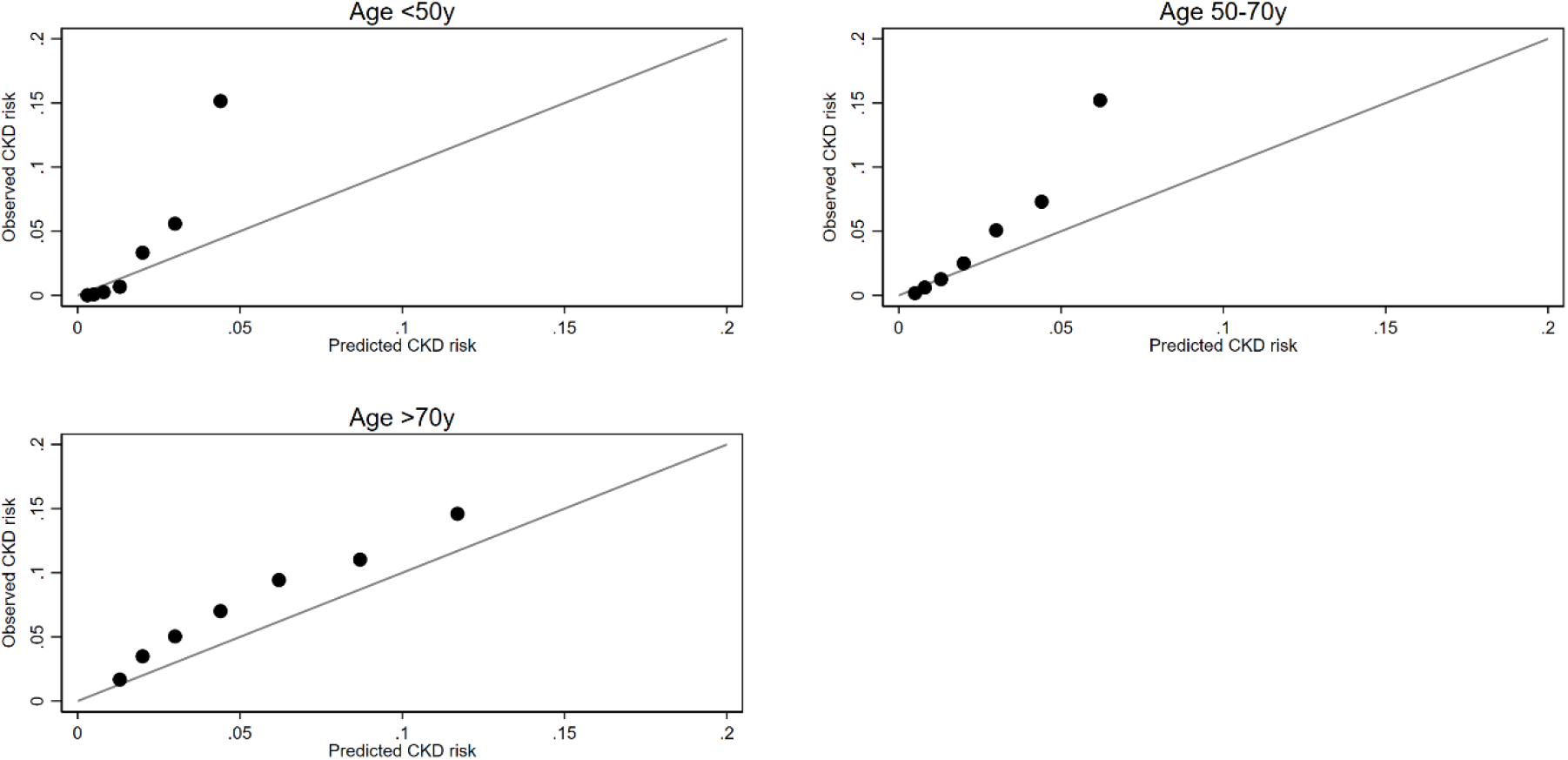
Observed versus predicted risk of CKD by age group. CKD chronic kidney disease

**Figure S2.**
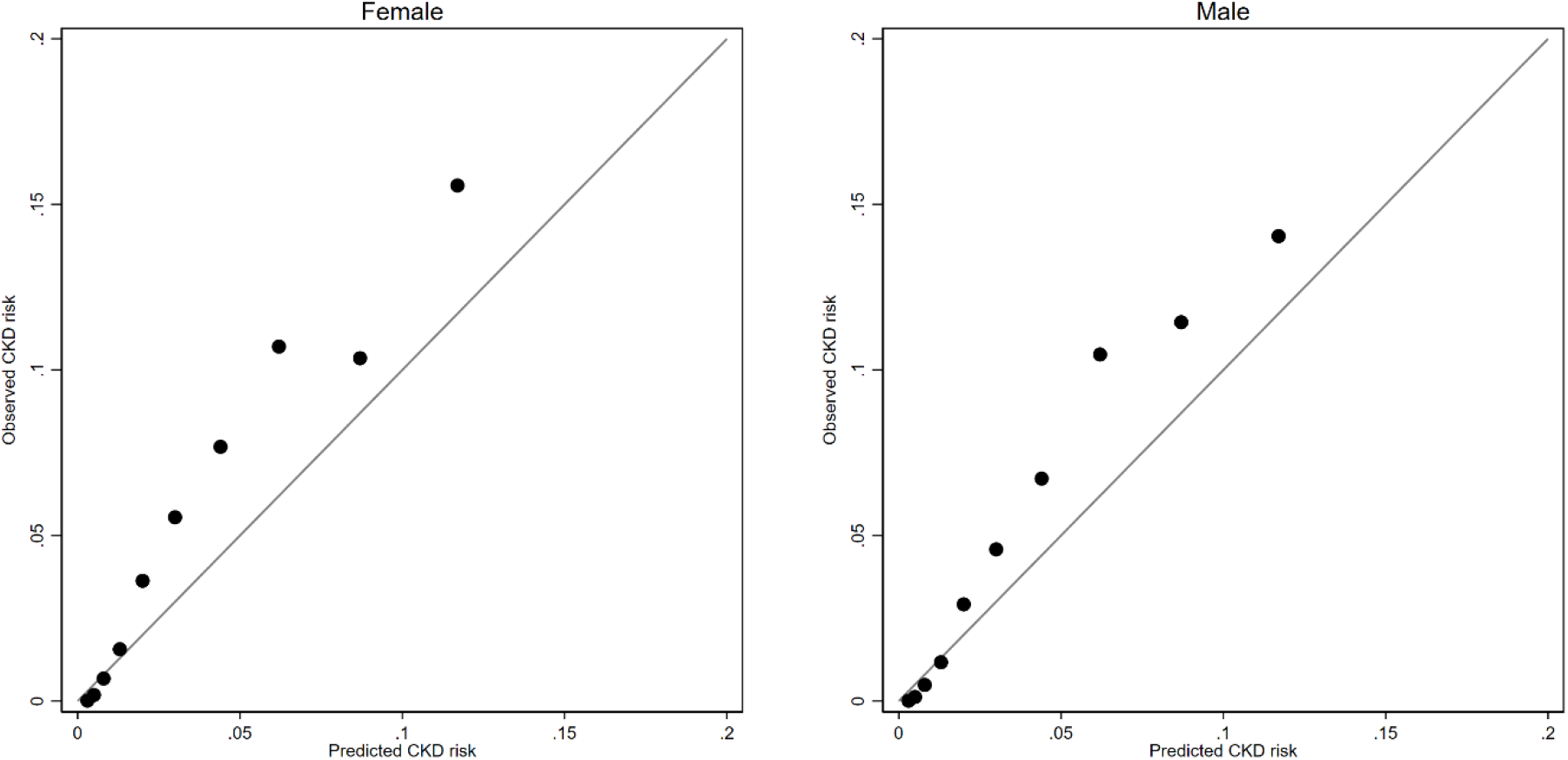
Observed versus predicted risk of CKD by sex. CKD chronic kidney disease

**Figure S3.**
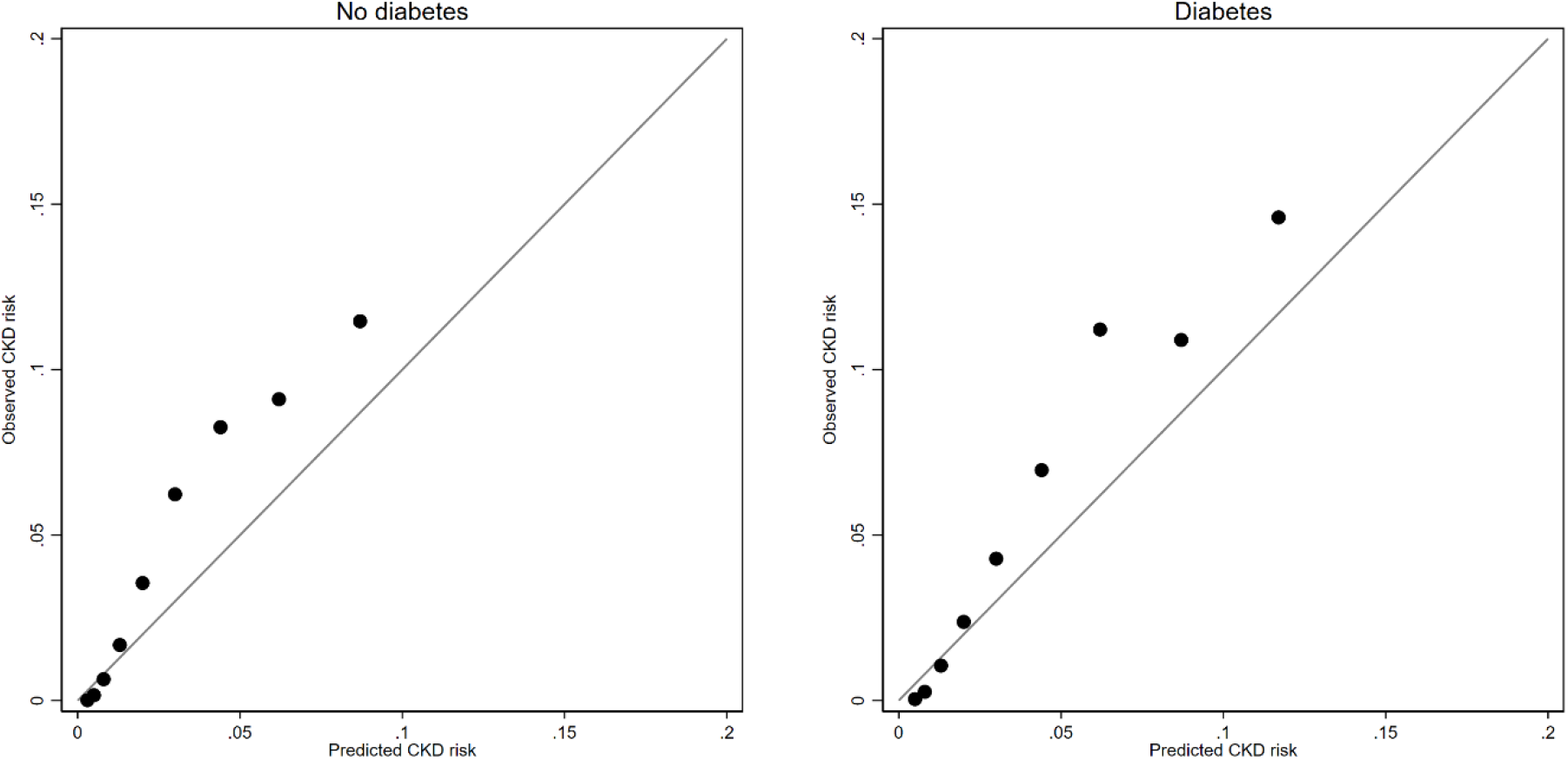
Observed versus predicted risk of CKD by diabetes status. CKD chronic kidney disease

## Notes

### Competing Interest Statement

The authors have declared no competing interest.

### Funding Statement

The study was funded by MTs David Freeze Chair in Health Services Research at the University of Calgary.

### Author Declarations

The institutional review boards at the Universities of Calgary and Alberta (REB16-1575/psite00000147) approved the study and waived the requirement for informed consent.

### Summary of Updates

The was an error in the abstract under findings. This sentence has been corrected: The final score had a maximum total of 9 points: age 50-70 years, moderate albuminuria, hypertension, diabetes and heart failure all received a single point, and age >70 years and severe albuminuria received three points.

